# A pragmatic randomized controlled trial of artificial intelligence (AI)-based predictive analytics monitoring for early detection of clinical deterioration

**DOI:** 10.1101/2025.01.20.25320838

**Authors:** Jessica Keim-Malpass, Sarah J Ratcliffe, Matthew T Clark, Katy N Krahn, Oliver J Monfredi, Susan Hamil, Gholamreza Yousevfand, Marieke K Jones, Ashley Nelson, Liza P Moorman, J. Randall Moorman, Jamieson M Bourque

## Abstract

**Background:** This pragmatic randomized controlled trial aimed to assess the effect of a passive display of artificial intelligence (AI)-based predictive analytics on hours free of clinical deterioration events among medical and surgical patients in an acute care cardiology medical-surgical ward.

**Methods:** 10,422 inpatient visits were randomly assigned by cluster to the intervention group of a display of risk trajectories or to a control group of usual medical care. The trial was undertaken on an 85-bed inpatient cardiology and cardiac surgery ward of an academic hospital with a substantial implementation and education plan. This was a passive display with no specific response mandated. The primary analysis compared events of clinical deterioration (death, emergent ICU transfer, emergent endotracheal intubation, cardiac arrest, or emergent surgery) and compared mortality 21 days after admission.

**Results:** Patients with a large spike in risk score had, on average, twice the length of hospital stay (6.8 compared to 3.4 days). There was no change in the primary outcome between groups. Among those who had a clinical event, there were more event-free hours in the intervention/display-on group compared to the standard-of-care/display-off, but this did not approach statistical significance. 11% of patients were transferred into or out of display beds, a censoring event removing them from the analysis, thereby undermining aspects of the randomized nature of the study.

**Conclusion:** Predictive analytics monitoring incorporating continuous cardiorespiratory monitoring and displays of risk trajectories coupled with an education plan did not improve patient outcomes or reduce deaths. While necessary to conduct the study, the pragmatic design allowed for significant movement towards intervention/displayed beds for sicker patients. Design considerations in the future must focus on understanding clinicians’ interpretation, care processes, and communication practices.

**Trial registration:** NCT04359641

## Introduction

Unanticipated clinical deterioration of hospital patients remains a pernicious problem that contributes to in-hospital morbidity and mortality.^1–4 5^ Often following the clinical deterioration event, clinicians who review these cases can recognize trends in vital signs, laboratory tests, and other electronic health record elements that, in retrospect, might have raised clinical suspicion of illness.^6^ For example, the coincidence of rising heart rate and falling blood pressure should suggest incipient shock, but often the changes in physiological parameters are subtle and insidious. Automated detection of such signatures of illness might inform clinicians of patients who could benefit from earlier assessment and proactive clinical action.^1,7^

Since its inception more than 20 years ago in the neonatal intensive care unit (ICU), ^8,9^ predictive analytics monitoring has been increasingly recognized for its potential to improve patient outcomes by early detection of deterioration. Artificial intelligence (AI) and machine learning methods support synthesizing unprocessed cardiorespiratory monitoring data, laboratory results and clinician-entered vital signs into computational models that can visually represent the estimation of the risk of clinical deterioration. ^10^ Since AI’s new and sometimes opaque methodologies lie at its core, predictive analytics monitoring has been held to high standards for transparency. For example, there are numerous guidelines and opinion pieces to inform the developer and the user.^11–13 14^

Incontrovertible proof of the benefit of predictive analytics monitoring in the form of large randomized clinical trials has been lacking. While a meta-analysis of artificial intelligence-based sepsis alerts showed an average mortality reduction of 30%, it included only six randomized trials with 2938 patients.^15^ To this list, we can add the original trial in this field, that of heart rate characteristics monitoring for neonatal sepsis in 3003 premature infants, which showed a 22% relative mortality reduction when risk estimates were displayed to clinicians. ^16^ Here, we extend the work in neonates to evaluate the impact of predictive analytics monitoring on events of clinical deterioration in a heterogeneous group of adults in acute care cardiac medical-surgical wards. We test the use of an AI-based risk analytic, Continuous Monitoring of Event Trajectories (CoMET, Nihon Kohden Digital Health Solutions, Irvine, CA), that visually displays risk estimates of multiple events of clinical deterioration every 15 minutes. We hypothesized that the passive display of a visual risk analytic would facilitate early identification of patients at risk for impending adverse outcomes and increase the hours free from clinical deterioration.

## Methods

### Study design

We conducted a cluster-randomized controlled trial (RCT) among patients admitted to 85-bed acute care cardiology medical-surgical wards. Eleven clusters were defined by room number, and patients were randomized to receive either the intervention (passive display of CoMET) or usual care (standard of care). Randomization began on January 4, 2021, and ended on October 4, 2022. Due to the pragmatic nature and replication of real-world patient contexts, all patients admitted to these floors during the study period were enrolled in the study. Clusters were re-randomized every 2 months following a Latin square design within each stratum to ensure a balanced treatment assignment. Full pre-registered study protocol details and additional randomization details can be found in ^17^, and the clinical trial was registered (NCT04359641).

The University of Virginia (UVA) IRB approved the study with a waiver of informed consent (IRB#22196). A Data Safety and Monitoring Board reviewed intercurrent results. The results are reported in accordance with the CONSORT-AI guidelines.^12^

### Patient care units and populations

The Medical-Cardiology unit predominantly comprised acute cardiology patients admitted with decompensated congestive heart failure, acute coronary syndrome, arrhythmias such as atrial fibrillation, and other cardiac diagnoses. The Medical-Surgical unit was historically the same as Medical-Cardiology. Immediately preceding the study, though, 11 of the 20 beds were repurposed for patients with solid organ transplants under the care of a dedicated transplant team. They differed from the patients and the practices of the cardiology and cardiac surgery beds. The baseline distributions of vital signs and laboratory test results, for example, differed, with a bias toward findings of chronic kidney and liver failure in the transplant patients. Due to the pragmatic nature of this study design in real-world contexts, they were left within the study population. The Cardio-Thoracic Surgery unit predominantly comprised patients awaiting discharge.

### Risk displays

We used the CoMET system to show two risk trajectories, calculated from previously published logistic regression machine learning analyses relating continuous cardiorespiratory monitoring parameters, nurse-charted vital signs, and laboratory test results to outcome events on these wards from 2013 to 2015. ^18^ The unit of measurement of the risk estimate is the fold-increase in the probability of the event in the next 24 hours compared to the average probability. Thus, a patient with a score of 1 has the average risk, a patient with a score of 2 has twice the average risk, and so on. The plot shows a comet-like icon for each patient with the bed number in the head; the tail is 3 hours long. Additional details on the model inputs and model performance, including excellent calibration, are in the following publication. ^19^

### Intervention implementation details

We followed principles derived from the prior implementation of predictive analytics monitoring in ICUs and non-ICUs at the UVA Hospital and a regional hospital.^20^ Each of the three units identified an interprofessional team of clinical champions to serve as super users, oversee implementation, and ensure staff education. Our team of educators (PIs, CoMET educators, clinical research coordinators (CRCs), and other study investigators) provided sessions with the clinical champions and other clinician groups in each unit before randomization and during the first quarter. We encouraged the integration of the CoMET scores into routine nurse vital sign assessments, rounding discussions, hand-off reports, and escalation protocols. Our recommended protocol for reporting and responding to changes in the CoMET scores included nurse notification of physicians or advanced practice providers of an increase in score by 2 units over a three-hour period and physician evaluation of the patient as appropriate. A CRC attended morning rounds on each unit on a rotating basis to show the display and answer questions about interpreting score trajectories. The CoMET scores flowed through to the electronic medical record every hour and were recorded within the vital sign flowsheet.^17^

### Study outcomes

The primary outcome was the number of hours free of clinical deterioration (adverse clinical events including: an emergent ICU transfer, emergent intubation, cardiac arrest, emergent surgery, and death) within 21 days of admission. A maximum score was 21 event-free days (504 hours), and patients who were successfully discharged from the hospital prior to the 21 days without an event were awarded all 504 hours. Patients who died during admission were counted as having 0 event-free days/0 hours. Patients with non-emergent ICU transfers, non-emergent/planned surgeries, or changes in bed assignments that caused a change in arm assignment were censored. Dedicated clinician CRCs adjudicated each case of clinical deterioration/study endpoint event. For any events that were ambiguous, the PIs of the study (JKM and JMB) blindly adjudicated outcomes through a weekly meeting.

### Statistical analysis

We used a generalized estimating equation with a zero-inflated negative binomial to analyze event-free hours and a Cox proportional hazards model to analyze time-until-first-critical events. The model handled the correlation of multiple hospital admissions from the same patient, within-unit clusters within the study design, and differences in the amount of time at risk for censored patients. An intent-to-treat analytic strategy was utilized where all randomized patients were included for the primary endpoint comparison between the intervention and usual care. As a post-hoc analysis, we also analyzed the primary outcome in an ‘at-risk’ cohort, or those patients who experienced a rise in 2 on either axis of the CoMET display during their hospitalization.

## Results

Over the 22 months of randomization, there were 10,422 patient visits within our study cohort (**Figure 1**). They were majority male, White, non-Hispanic, and on average 65 years of age with relatively low hospital length of stays (**Table 1**). Model performance is reported separately ^19^, but overall the predictive models performed well and outperformed a common early warning score, qSOFA.

**Table 1:**
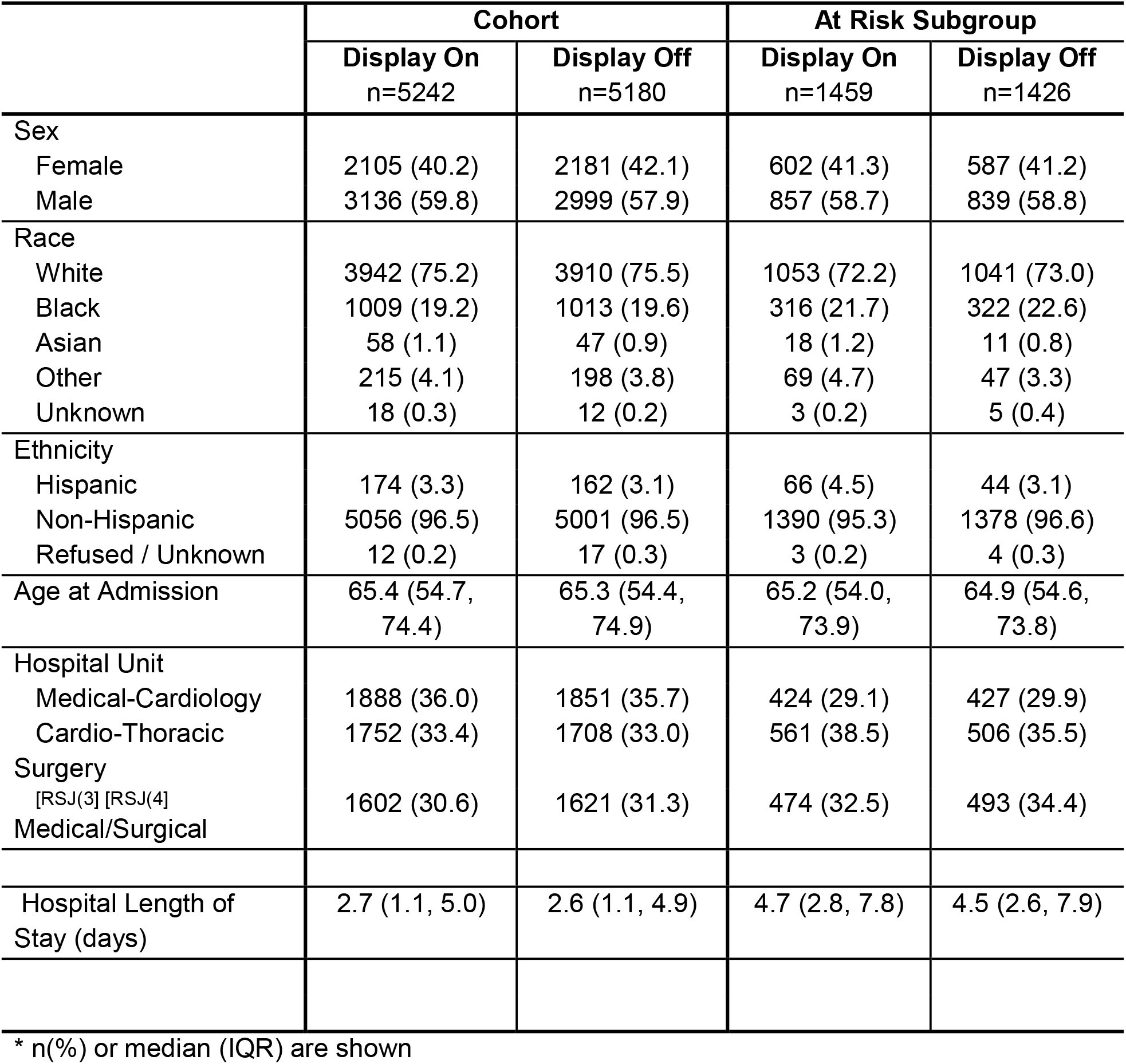
Admission Characteristics of the Cohort and At-Risk Subgroup by Arm.

**Figure 1.**
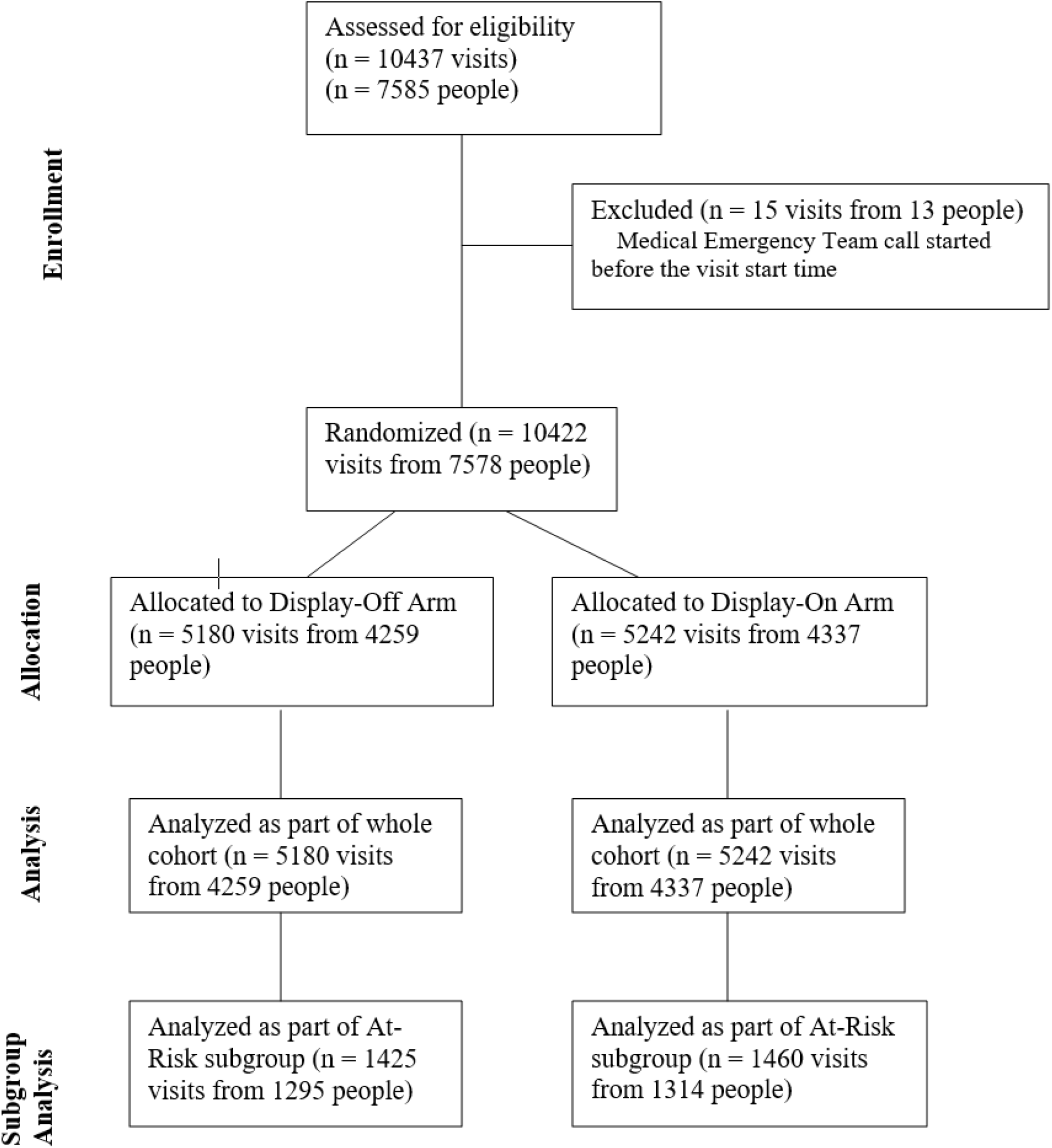
CONSORT randomization information

The vast majority of participants were discharged successfully and had the maximum amount of event-free hours (**Table 2**). There was no statistically significant difference in the primary outcome by study arm in both the full cohort and those at risk for clinical deterioration. Among those who had a clinical event, there were more event-free hours in the intervention/display-on group compared to the standard-of-care/display-off, but this did not approach statistical significance. Censoring events and components meeting the endpoint of clinical deterioration are found in Table 2. The estimated marginal means from the negative binomial generalized estimating equation are found in **Figure 2**.

**Table 2:** Study Outcomes for the Cohort and At Risk Subjects.

|  | Cohort |  |  | At-Risk Subgroup |  |  |
| --- | --- | --- | --- | --- | --- | --- |
|  | Display On<br>n=5242 | Display Off<br>n=5180 | IRR/OR<br>(95%<br>CI) | Display On<br>n=1459 | Display Off<br>n=1426 | IRR/OR<br>(95% CI) |
| **Primary Outcome:** |  |  |  |  |  |  |
| Event-Free Hours | 504<br>(0-504) | 504<br>(0-504) | 0.995<br>(0.88-1.12) | 504<br>(0-504) | 504<br>(0-504) | 1.07<br>(0.91-1.26) |
| **Outcome censored due to:** |  |  |  |  |  |  |
| N/A | 4461 (85.1) | 4411<br>(85.1) |  | 1204<br>(82.5) | 1168<br>(81.9) |  |
| Bed change | 617 (11.8) | 595<br>(11.5) |  | 195<br>(13.4) | 197<br>(13.8) |  |
| Re-randomization | 164 (3.1) | 174 (3.4) |  | 60 (4.1) | 61 (4.3) |  |
| **Components of Primary Outcome:** |  |  |  |  |  |  |
| Had any Event | 282 (5.4) | 271 (5.2) | 1.01<br>(0.85-1.20) | 179<br>(12.3) | 171<br>(12.0) | 1.05<br>(0.84-1.32) |
| Event-Free Hours in Patients with an Event<sup>1</sup> | 100.6<br>(0-500) | 95.0<br>(0-499) | 0.99<br>(0.88-1.12) | 142.8 (0-500) | 90.4 (0-498) | 1.07<br>(0.91-1.26) |
| Death | 57 (1.1) | 42 (0.8) | 1.42<br>(0.93-2.16) | 37 (2.5) | 28 (2.0) | 1.45<br>(0.86-2.46) |
| Emergent ICU Admission | 224 (4.3) | 233 (4.5) | 0.94<br>(0.78-1.13) | 151<br>(10.3) | 150<br>(10.6) | 0.99<br>(0.90-1.27) |
| Emergent Intubation | 65 (1.2) | 48 (0.9) | 1.31<br>(0.89-1.92) | 38 (2.6) | 30 (2.1) | 1.29<br>(0.78-2.13) |
| Cardiac arrest followed by ICU/Death | 22 (0.4) | 25 (0.5) | 0.84<br>(0.45-1.54) | 12 (0.8) | 14 (1.0) | 0.96<br>(0.42-2.19) |
| Emergent transfer for Surgery | 22 (0.4) | 16 (0.3) | 1.34<br>(0.68-2.61) | 11 (0.8) | 8 (0.6) | 1.67<br>(0.66-4.21) |
| First Event that Occurred |  |  |  |  |  |  |
| Death | 21 (0.4) | 16 (0.3) |  | 14 (1.0) | 10 (0.7) |  |
|  | Display On<br>n=5242 | Display Off<br>n=5180 | IRR/OR<br>(95% CI) | Display On<br>n=1459 | Display Off<br>n=1426 | IRR/OR<br>(95% CI) |
| **Primary Outcome:** |  |  |  |  |  |  |
| Emergent ICU | 198 (3.8) | 201 (3.9) |  | 133 (9.1) | 131 (9.3) |  |
| Emergent Intubation | 24 (0.5) | 16 (0.3) |  | 11 (0.8) | 9 (0.6) |  |
| Cardiac arrest followed by ICU/Death | 18 (0.3) | 22 (0.4) |  | 10 (0.7) | 11 (0.8) |  |
| Surgery | 21 (0.4) | 16 (0.3) |  | 11 (0.8) | 8 (0.6) |  |

<sup>1</sup> N = those who had an event
n(%) or median (inter-quartile range). IRR/ORs for display-on versus display-off using negative binomial or logistic regression adjusted for study design.

**Figure 2:**
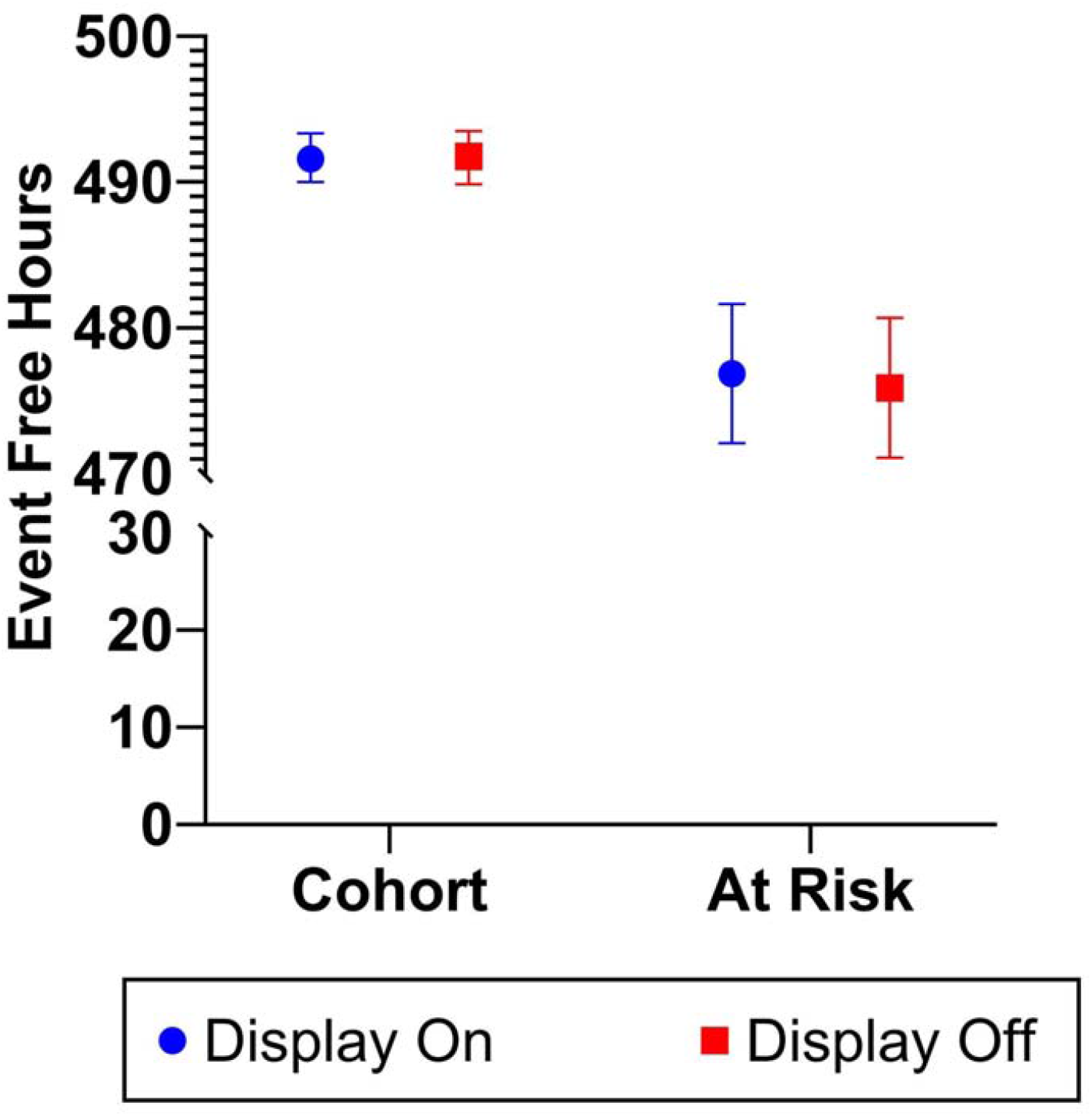
Estimated marginal mean (and 95% CI) event-free hours by arm and population

Bed movement was common within this RCT (n=382 went from usual care to CoMET display, and n=419 went from CoMET display to usual care). In a *post-hoc* analysis, we found higher relative risk of clinical deterioration among those transferred from a non-display to a display bed. This finding indicates that clinicians made decisions that undermined the cluster RCT design by preferentially placing sicker patients into the CoMET display beds, presumably to obtain additional information about their underlying physiological instability. (**Figure 3**).

**Figure 3:**
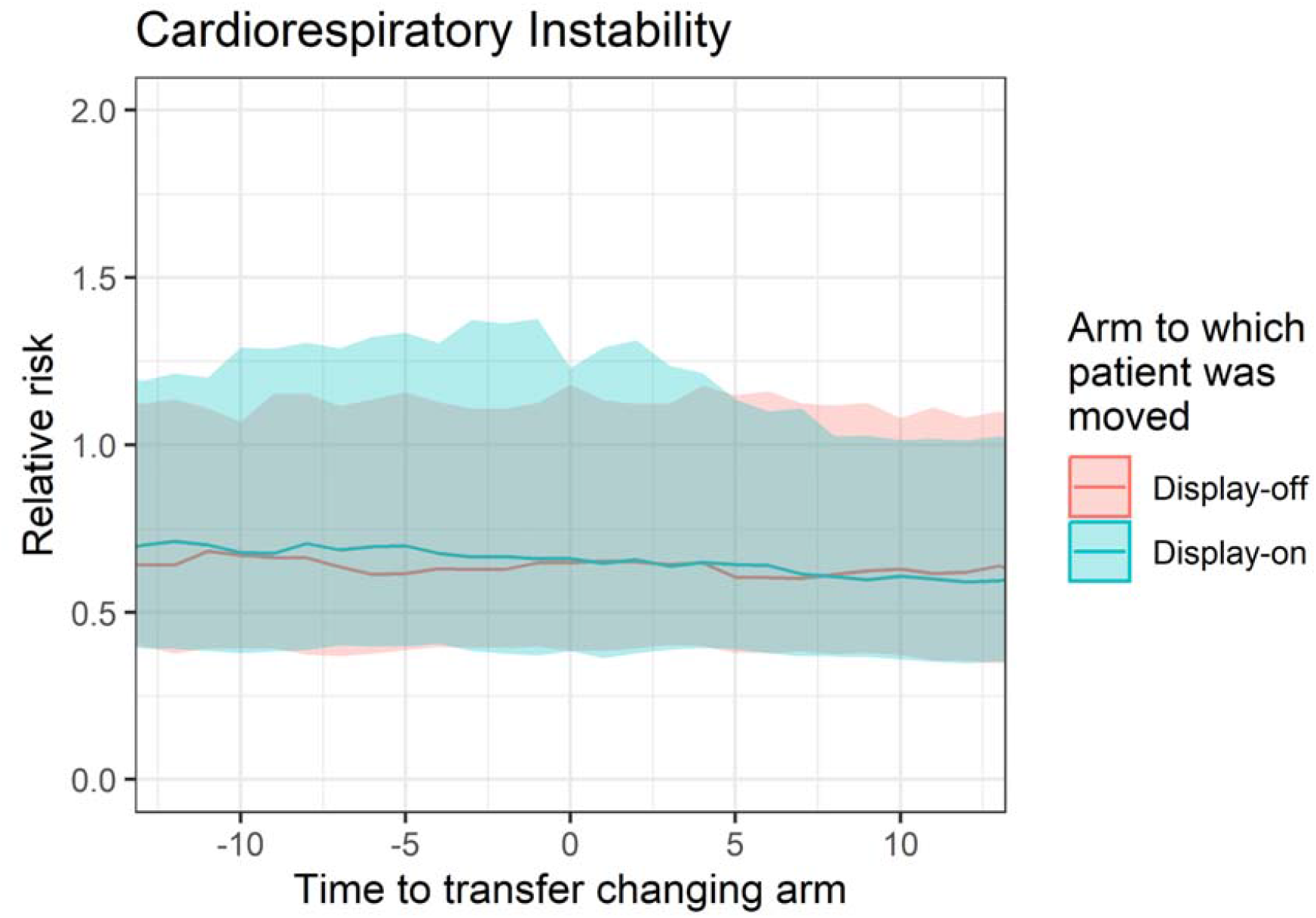
Time zero is transfer from one arm to another. The line is the median and the shaded region is the IQR. Blue are those patients transferred from a display-off to display-on bed and their overall relative risk of clinical deterioration is higher, representing a sicker population.

## Discussion

We conducted a large randomized controlled trial to test the impact of predictive analytics monitoring on cardiology and cardiac surgery patients in in-patient acute care wards. Our major findings are that a display of the trajectories of the risk of imminent deterioration without alerts was not associated with an increase in hours free of clinical deterioration. We found that there was an increase in relocating sicker patients from usual care to CoMET display-on, thereby distorting the RCT design. We note this study was undertaken during the early stages of the COVID-19 pandemic in our area, which disrupted staffing and resource utilization^10^ and was associated with a lower event rate than our pre-COVID baseline.

We justify using continuous cardiorespiratory monitoring in the predictive models because of the causality leakage of clinician-ordered tests. For example, the presence of a serum lactate measurement in the chart bespeaks clinical suspicion of sepsis. It excels as a predictor, but not because it detects changes before clinicians. Likewise, the use of pressors, fluid administration, and blood cultures as predictors rather than endpoints allows causality leakage, artificially inflating model performance. Kamran and coworkers recently showed that the Epic Sepsis Model had no predictive capability at all when assessed prior to any clinician suspicion or action, that is, before fluids, antibiotics, and lactate tests. ^21^

We justify the randomized clinical trial approach to the study of AI-based risk analytics. We find irreducible uncertainty in other trial designs and results, even our own demonstration of a 50% reduction in septic shock in a surgical ICU after the introduction of CoMET trained for urgent unplanned intubation for acute respiratory failure and hemorrhage requiring large transfusion. ^22^ While a control medical ICU without CoMET monitoring had no consistent benefit over the same time period, the absence of randomization has allowed skepticism of the before-and-after study results. We especially endorse the simple and pragmatic nature of the current trial -we enrolled everyone to mimic complex real-world acute care environments and used important endpoints to clinicians and patients.

Other studies have used non-randomized designs. For example, Winslow, Edelson, Churpek, and coworkers found reduced mortality throughout the hospital after initiating their early warning system. ^23^ The magnitude was substantial but was the same even for patients with low risks who were never alerted. Adams, Henry, Saria, and coworkers used a machine learning-based sepsis early warning system that included mechanical ventilation, lactate, and vasopressors as sepsis predictors and placed passive alerts in the EHR, but only after verifiable symptoms were present. They found reduced mortality in the 1.2% of patients who met retrospective sepsis criteria and whose alerts were acknowledged within three hours but did not present outcomes of the other patients. ^24,25^ Groups led by Shimabukuro ^26^ and Watkinson ^27^ performed small randomized trials with divergent results. Escobar, Liu and coworkers used a non-randomized stepped wedge approach to show a convincing mortality reduction with an EHR-based early warning score in 19 hospitals. ^28^ They employed an informed intermediary to receive alerts and to review the clinical record before alerting bedside clinicians.

Cluster randomization was chosen because clinicians needed to easily recognize who was in the intervention group. We did not want to add additional burden to the complexity of the acute care environment and did not want to falsely reassure a clinician who did not see a rising score to assume the patient was in the intervention group. Although we have previously tested the positive predictive value of creating a threshold-based alert from CoMET ^1^, we did not want to add to clinician burden or create additional markers of alarm fatigue and decided to offer suggestions for response without an active alert strategy.

We approached education and implementation of novel analytics in real-world environments within the context of the COVID-19 pandemic and wanted to minimize any contribution to clinician burnout. ^29^ Implementation considerations focused on promoting trust, transparency, integration with workflow, prolonged stakeholder engagement, tailored education embedded within hospital norms, supporting actionability through integration within clinical care, and sustainability through ongoing supportive interactions. ^29^

AI clinical trials call for an understanding of new and separate design elements not previously contemplated. There are several elements of this type of intervention such asthe use of passive displays that warrant further study and have RCT design implications when studying AI-based predictive analytics: the proactive nature of the clinical action response, the rarity of the event of interest, and the complexity of implementation within evolving acute care environments. While the pragmatic design allowed for implementation in real-world environments, this also meant there was heterogeneity in patient populations, unit work environments, and clinician responses. The real-world context of clinical trials such as this implies that the implementation of AI-based risk analytics must also be adaptive to ensure long-term integration and practice flexibility across different work environments. Understanding the cognitive interpretation of scores in real-time, the response through diverse care processes, and communication patterns involving AI scores preceding clinical events is an area in particular need of study. In hindsight, hours free of events of clinical deterioration may not have been the most sensitive primary outcome.

Outcomes specific to the at-risk population and subsequent intensive care unit trajectories may have been more appropriate, given the overall rarity of our event.

## Data Availability

All data produced in the present study are available upon reasonable request to the authors

